# Mammography utilization and male breast cancer in *BRCA1/2* carriers

**DOI:** 10.64898/2026.09.22.26363613

**Authors:** Meredith Y. Davies, Maliha Tayeb, Emily Scarrow, Jasmine Phun, Evelyn Milavsky, Caitlin Orr, Roukiatou Sore, David Y. Lu, Heather Symecko, Sarah Ehsan, Heena Desai, Ryan Hausler, Bradley Wubbenhorst, Abigail Doucette, Peter E. Gabriel, Katherine L. Nathanson, Christine E. Edmonds, Kyle W. Robinson, Michael J. Kelley, Isla P. Garraway, Leah L. Zullig, Erin M. Bayley, Haley A. Moss, Martin W. Schoen, Anne Marie McCarthy, Susan M. Domchek, Jennifer Q. Zhang, Kara N. Maxwell

## Abstract

**Purpose:** Men with pathogenic *BRCA1* and *BRCA2* variants (*BRCA+*) have an elevated lifetime risk of male breast cancer (MBC) yet data regarding mammography in this population remain limited.

**Methods:** We performed a retrospective cohort study of *BRCA*+ assigned male at birth individuals (males) at Penn (1/1/2008-5/1/2024) and the Veterans Administration (VA, 10/1/2002-1/1/2026) for mammography uptake. Mammography results and MBC characteristics were compared between *BRCA*+ and *BRCA*-males.

**Results:** Among 810 *BRCA+* males, 19% had a documented mammogram for screening or symptoms. Among 562 *BRCA*+ males age-eligible for mammogram without prior MBC, most (63%) had documentation of a mammography-related discussion and 16% completed mammogram, of whom 25% were *BRCA1*+ and 75% were *BRCA2+*. Among mammograms obtained for screening of 97 *BRCA+* males, there were few BI-RADS 4/5 (N=1, 1%) and no MBC was detected. Symptom-indicated mammograms in 59 *BRCA*+ compared to 228 *BRCA*-males yielded BI-RADS 4/5 findings in 35% versus 25%, p<0.0001, and MBC diagnoses in 29% versus 21%, p=0.219. The positive predictive value (PPV) of BI-RADS 4/5 lesions was similar in *BRCA+* vs *BRCA-*males (71% vs 74% and 100% vs 93%, respectively). Among 22 *BRCA*+ and 57 *BRCA*-MBC, 55% and 42% were node positive and 82% and 86% were ER+Her2-, respectively.

**Conclusion:** Screening mammography utilization among *BRCA*+ males was low despite elevated hereditary risk. Regardless of *BRCA* status, nearly all MBC were diagnosed based on symptoms, and a high proportion of MBC were diagnosed at advanced stages. Our findings identify a critical need for both improved identification of males at high risk for breast cancer and improved screening practices of high-risk men.

## Introduction

Male breast cancer (MBC) is a rare disease, accounting for approximately 1% of all breast cancers worldwide, with an estimated lifetime risk of 0.1% in the general male population^1–4^. Men are often diagnosed at older ages and with more advanced-stage disease compared to women, contributing to worse outcomes^1^. Histologically, most MBCs are invasive ductal carcinomas (IDC) with high rates of hormone receptor (HR) positivity, particularly estrogen receptor (ER) expression^4^. Management strategies for MBC are largely extrapolated from female breast cancer studies, and dedicated screening and treatment guidelines remain limited^2,5–7^.

Hereditary cancer syndromes, particularly pathogenic germline variants (PGV) in *BRCA1* and *BRCA2* (*BRCA*+) are identified in up to 14% of MBC cases^8–11^. *BRCA2* carriers have a 4%-10% estimated lifetime risk of MBC, and *BRCA1* mutations confer a lower but still elevated lifetime risk of approximately 1%-5%^9,12–16^. *BRCA1*+ women with BC are more likely to be diagnosed at younger ages with more advanced tumors^17,18^, however, it is unknown if there are worse clinicopathological features or outcomes among *BRCA*+ males. Finally, beyond *BRCA1/2*, other genetic and non-genetic risk factors for MBC remain under investigation.

Despite elevated risks of MBC, screening recommendations for *BRCA*+ men are poorly defined^19^. Current National Comprehensive Cancer Network (NCCN) guidelines (version 2.2026) for *BRCA+* carriers include annual clinical breast examinations beginning at age 35 with consideration of screening mammography starting at age 50 or 10 years prior to the earliest known male breast cancer in the family (whichever comes first), especially for those carrying a *BRCA2* PGV ^20,21^. Consideration of mammography was added to NCCN guidelines in February 2023 (version 2.2023). However, these recommendations are based on limited evidence and prospective studies evaluating mammography in men are lacking^2,9,22^. Existing data suggests that mammography performs well in the male breast, with reported sensitivities exceeding 90%^23^. In high-risk men, cancer detection rates have been reported to be comparable to those observed in female screening populations with one study reporting detection rates of 4.9 per 1,000 examinations in high risk men versus 5.4 per 1,000 in average-risk women^24^.

Mammographic interpretation is standardized using the American College of Radiology Breast Imaging Reporting and Data System (BI-RADS). In women, BI-RADS category 4 lesions are considered suspicious abnormalities with a broad estimated malignancy risk (7-97%) and are subdivided into 4A, 4B, and 4C based on increasing likelihood of malignancy^25–27^. In female cohorts, the positive predictive value (PPV) of BI-RADS 4 lesions increases substantially across subcategories, with reported PPVs ranging from 7.6% for BI-RADS 4A to 69.3% for BI-RADS 4C^26^. Although BI-RADS classification is central to breast imaging decision-making in women, its predictive performance in men is poorly characterized^22,24,28^.

Given the increasing identification of men with hereditary cancer predisposition through genetic testing, an improved understanding of breast imaging utilization and outcomes in this population is needed. Data describing mammographic findings, BI-RADS categorization, and diagnostic performance in men remain limited. Further investigation into the role of mammography in high-risk male populations may help refine screening strategies and support earlier detection of male breast cancer.

## Methods

### Study Cohorts

Two cohorts were created, the first a Mammography Decision-Making Cohort to evaluate decision making among *BRCA+* males eligible for consideration of mammogram (**Figure 1**). A second a Mammography/MBC cohort investigating screening outcomes among males undergoing mammography. Both cohorts were developed at Penn Medicine and the Veterans Health Administration and combined for analysis. Inclusion Criteria for the Mammography Decision-Making Cohort were assigned male at birth (AMAB), age ≥50 years, and carrier of *BRCA1/2* PGV. Inclusion criteria for the Mammography/MBC Cohort were AMAB, documentation of completed mammogram in medical record, and germline genetic testing completed.

**Figure 1:**
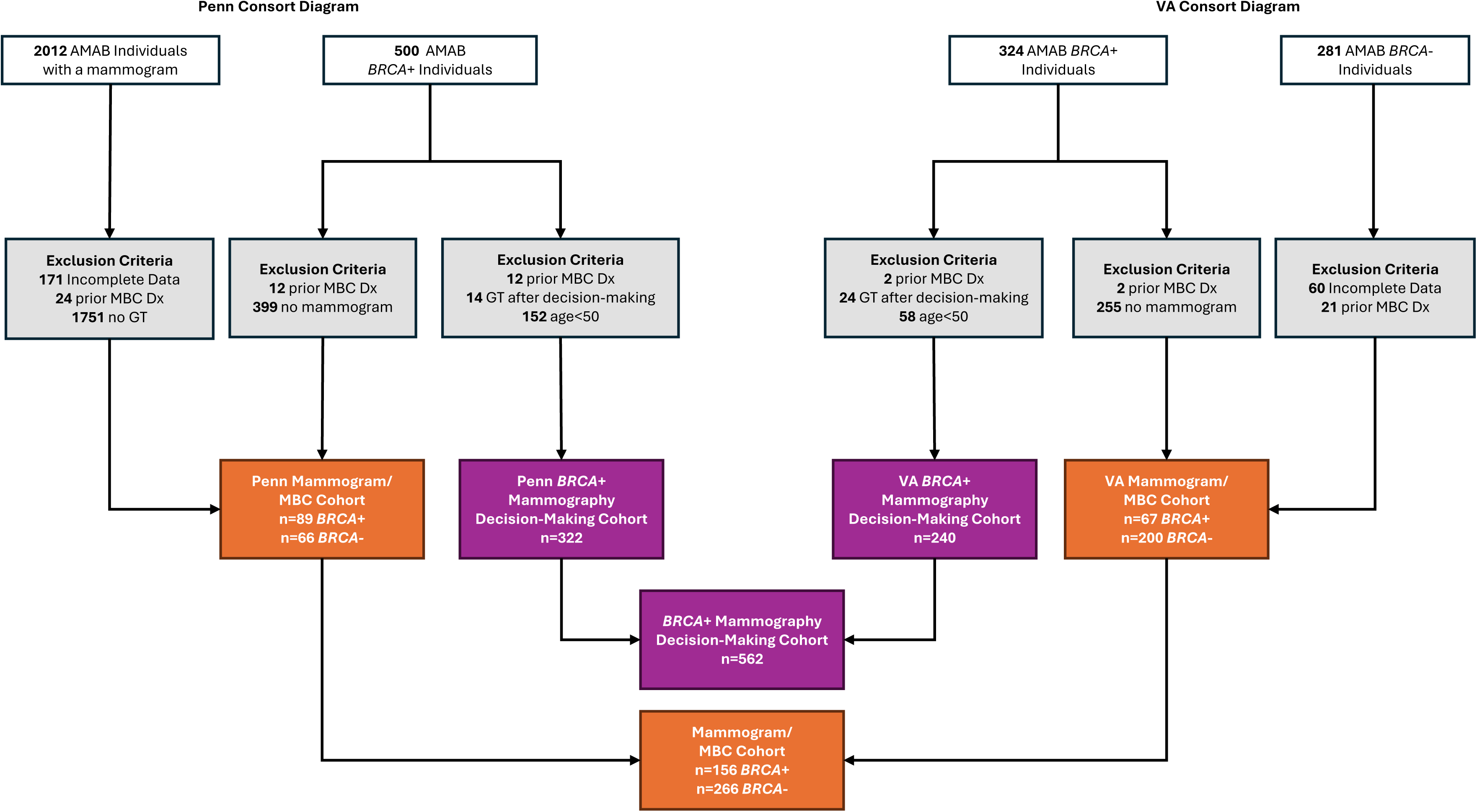
Consort diagram of *BRCA*+ and *BRCA*-AMAB individuals (males) identified at Penn Medicine and Veterans Health Administration. Males with a confirmed *BRCA1/2* PGV prior to mammography, no confirmed prior MBC diagnosis and age>50 comprised the *BRCA*+ Mammography Decision Making Cohort. Males with no prior MBC diagnosis, complete mammogram data, and germline genetic testing comprised the Mammogram/MBC cohort. Abbreviations: AMAB, assigned male at birth; GT, genetic testing; MBC Dx, male breast cancer diagnosis; PGV, pathogenic germline variant

### Penn Medicine Study Cohort

Institutional review board (IRB) approval was obtained to abstract patient information from the Penn Medicine electronic health record (IRB #854011, IRB #842712). A retrospective review of medical records of 500 *BRCA1+* and *BRCA2*+ (*BRCA*+) AMAB individuals – herein referred to as “males” – identified at Penn Medicine between January 1, 2008 to May 1, 2024 was conducted (**Figure 1)**. Germline carrier status of each individual was manually verified in the electronic health record. To create the Penn *BRCA*+ Mammography Decision-Making Cohort, 12 patients were excluded due to a diagnosis of MBC prior to the first mammogram on record, 14 due to having genetic testing after their mammogram, and 152 due to being age <50, making a total of 322 *BRCA*+ males. To create the Penn Mammogram/MBC Cohort, 399 of 500 males were excluded due to no reported mammography and 12 were excluded due a diagnosis of MBC prior to the first mammogram on record, for a total of 89 included *BRCA*+ males who had at least one mammogram. For comparison, 2012 males who had completed at least one mammogram were obtained from the Epic Mammography Quality Standards Act (MQSA) query. Males were excluded if mammography data was incomplete (for example missing indication or missing report in the health record, n=171), for a prior MBC diagnosis (n=24), and/or no prior germline genetic testing by manual review of the medical record (n=1751), for a total of 66 *BRCA*-males with at least one mammogram (Mammogram/MBC Cohort, Penn *BRCA*-).

### Veterans Health Administration Study Cohort

At the VA, IRB approval (#1742695) was obtained and clinical germline genetic testing data was extracted from the Veterans Affairs Informatics and Computing Infrastructure (VINCI) data warehouse and manual chart review (including records for care conducted at non-VA facilities referred to as community care records) identified 324 *BRCA*+ males between October 1, 2002 to January 1, 2026. To create the VA *BRCA*+ Mammography Decision-Making Cohort, two patients were excluded from the cohort due to a diagnosis of MBC prior to the first mammogram on record, 24 due to having genetic testing after their mammogram, and 58 due to being age<50, making a total of 240 *BRCA*+ males (**Figure 1**). To create the VA Mammogram/MBC Cohort, 255 were excluded due to no reported mammography and two were excluded due a diagnosis of MBC prior to the first mammogram on record, for a total of 67 included *BRCA*+ males. For comparison, 281 *BRCA-*male veterans, with at least one mammogram completed within the VA system were identified from VINCI. Males were excluded if mammography data was incomplete (n=60) or for a prior MBC diagnosis (n=21), for a total of 200 males analyzed in the *BRCA-*cohort (Mammogram/MBC Cohort, VA *BRCA*-).

### Study Variables

For the *BRCA*+ Mammography Decision-Making Cohort (n=562), mammography discussion data was obtained from review of clinical notes. Discussions were summarized into five categories: no discussion documented, the option of mammogram was discussed by a genetic counselor but no ordering provider discussion, shared-decision making discussion against mammogram, shared-decision making for a mammogram resulting in an order being placed but imaging not completed, and completed mammogram.

For the Mammogram/MBC Cohort (n=156 *BRCA*+, n=266 *BRCA*-), data collected included demographic characteristics, mammography findings, and MBC outcomes prior to 10/1/25 at Penn Medicine and 8/1/26 at VA. Mammography variables included age at first mammogram, BI-RADS assessment category, breast density, and indication for imaging (screening-indicated vs. symptom-indicated). Screening-indicated imaging was defined as mammography performed due to genetic risk, gynecomastia in the absence of other symptoms, incidental findings on other imaging, or family history in the setting of negative genetic testing. Screening-indicated imaging included diagnostic mammograms ordered in asymptomatic males. Symptom-indicated imaging was defined as mammography completed due to findings such as palpable masses, nipple changes or discharge, breast pain, swelling, or rash. For patients diagnosed with MBC, age at diagnosis, hormone receptor status, stage at diagnosis, and Oncotype DX genomic test recurrence scores were collected. Patients were followed for outcomes of disease progression including diagnosis of metastatic disease and contralateral breast cancer for 3-10 years (Penn) and 0.8-16 years (VA) from their date of MBC diagnosis.

### Statistical Analysis

Statistical analyses were performed using GraphPad Prism 10.6.1. Continuous variables, including age at initial mammogram and age at MBC diagnosis, were compared using Student’s t-test. Fisher’s exact test was used to compare categorical variables. Chi-square analyses were performed to compare categorical variable distributions, with Fischer’s exact tests used in comparisons with small cell sizes (N<5). All tests were two-sided, and a p value <0.05 was considered statistically significant.

## Results

### Screening mammography decision making in BRCA+ males

We identified 810 *BRCA*+ males without a prior MBC diagnosis across two healthcare systems. To investigate mammography uptake among this population, we created a *BRCA*+ Mammography Decision-Making Cohort, which included 562 males without a prior MBC diagnosis, age ≥50 and therefore eligible for mammography and had knowledge of their *BRCA* status prior to decision-making (**Figure 1**). Among these 562 *BRCA*+ males, 91 (16%) completed a mammogram. (**Figure 2, Supplementary Table 1**). Of the *BRCA*+ males who did not have a mammogram, 107 (19%) had documentation that a genetic counselor discussed annual mammography, but no subsequent discussion with an ordering provider was recorded. Documentation of shared decision-making (SDM) with an ordering provider resulting in a decision not to pursue mammography was found in 134 (24%) of males, and mammography was ordered but not completed in 23 (4%). Finally, no discussion regarding mammography was found for 207 (37%) eligible *BRCA*+ males. We investigated whether mammography completion rate varied by *BRCA1* versus *BRCA2* status in males who either completed a mammogram or had a SDM discussion but no mammogram completed. Mammography completion rate was similar in *BRCA1*+ versus *BRCA2*+ males, 23/196 (12%) versus 68/363 (19%), p=0.050 (**Figure 2**).

**Figure 2:**
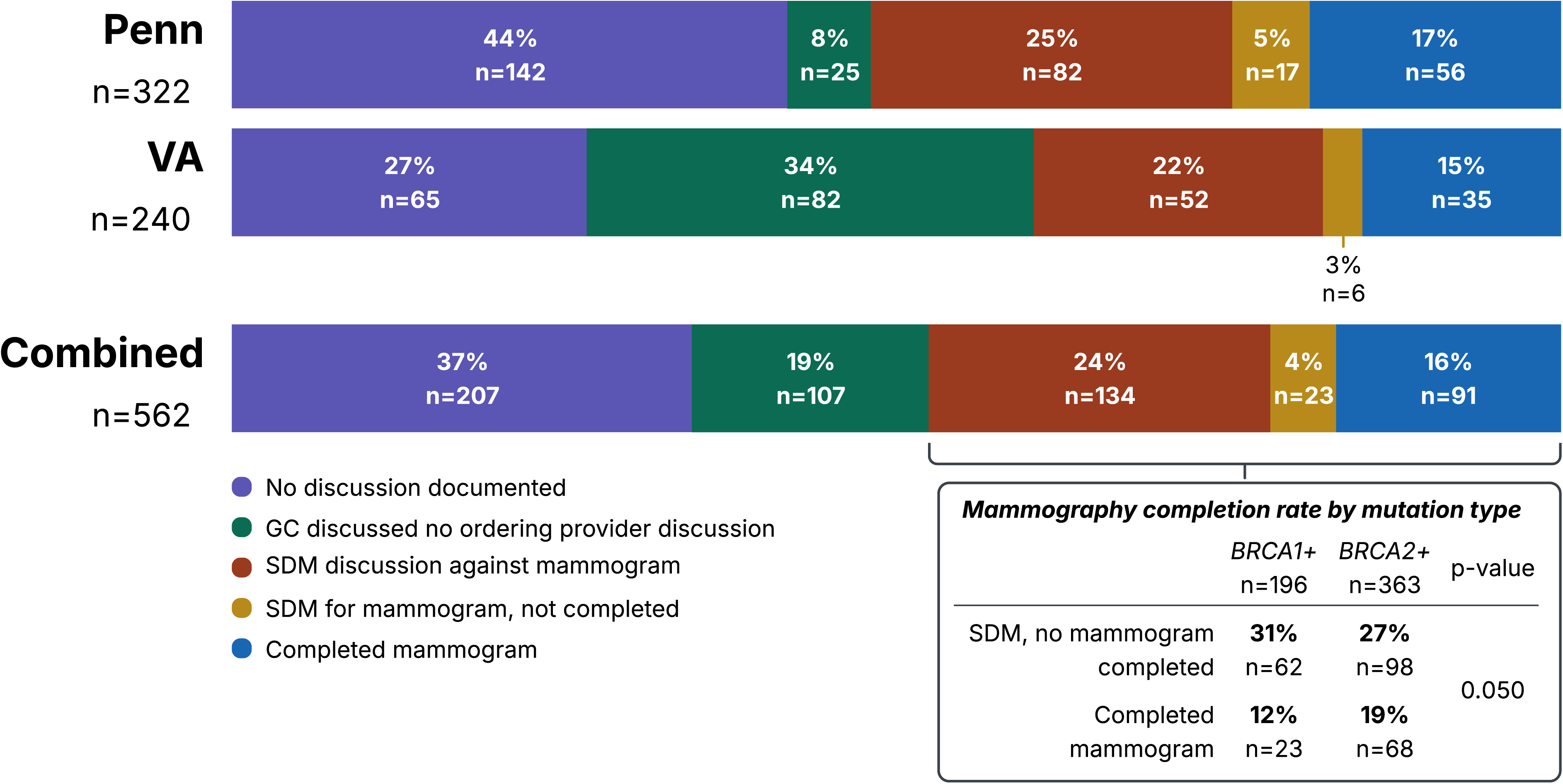
Summary of documented discussions in medical record regarding mammography in *BRCA+* males 50 or older at Penn Medicine, VA and in the Combined Cohort. Mammography completion rate stratified by mutation status for males with SDM against mammogram, SDM for mammogram but not completed and completed mammogram. Abbreviations: GC, genetic counselor; SDM, shared decision making

### Mammography findings and breast cancer diagnoses in males

We next investigated mammography findings and MBC diagnoses in the entire cohort of *BRCA*+ males that underwent mammogram, including males age <50 years and those who did not know their genetic status at the time of mammogram (n=156) and compared to 266 *BRCA-*males (Mammogram/MBC Cohort) (**Table 1, Supplemental Table 3**). The *BRCA*+ cohort was comprised of 128/156 (82%) self-identified White and 15/156 (10%) self-identified Black or African American males. In comparison, the *BRCA-*cohort was more racially diverse with 183/266 (69%) self-identified White males and 67/266 (25%) self-identified Black or African American males (p<0.001).

**Table 1:**
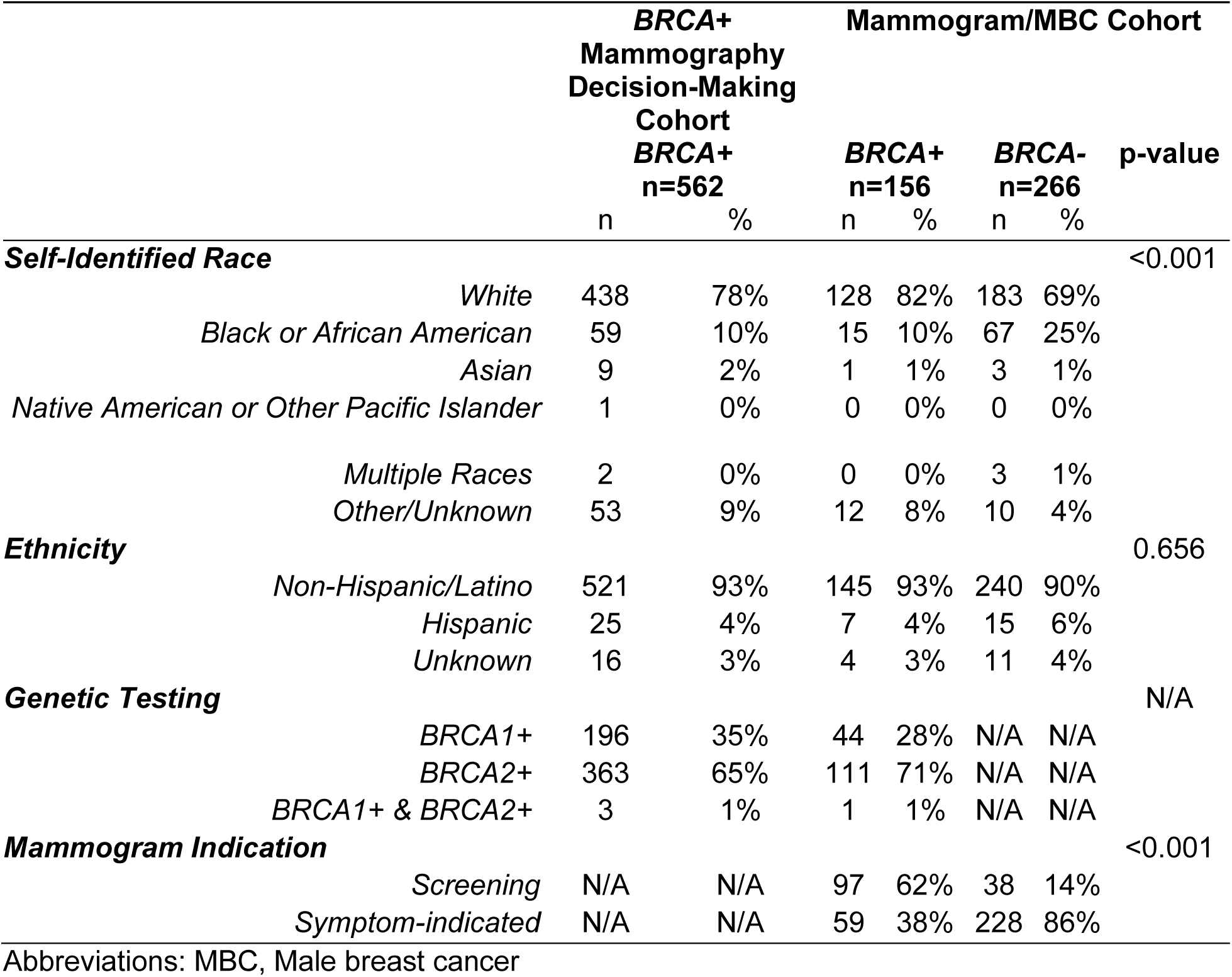
Demographics of Cohorts.

Among the 156 *BRCA*+ males, the reason for performing the first documented mammogram on record was primarily for screening indications (97/156, 62%), compared to symptoms (**Table 1**). Among 266 *BRCA*-males, imaging was completed predominantly for symptoms (228/266, 86%, p<0.001). When stratified by indication, *BRCA*-males undergoing screening mammography for gynecomastia in absence of other symptoms, incidental findings on other imaging, or family history in setting of negative genetic testing had higher rates of BI-RADS 4/5 findings than *BRCA*+ males (6/38, 16% vs 1/97, 1%, p=0.337). Six MBC (16%) were diagnosed on screening-indicated mammograms in *BRCA*-males; whereas no MBC were diagnosed in *BRCA*+ males undergoing screening mammography (**Table 2, Supplemental Table 4**). Among patients undergoing symptom-indicated mammography, rates of BI-RADS 4/5 findings were higher in *BRCA*+ versus *BRCA*-males (21/59, 35% vs 58/228, 25%, p<0.001), but the MBC diagnosis rates were not statistically different between *BRCA*+ and *BRCA*-males (17/58, 29% vs 47/228, 21%, p=0.219).

**Table 2:** Mammography results in *BRCA*+ versus *BRCA*-males.

|  | Screening Indicated |  |  |  |  | Symptom Indicated |  |  |  |  |
| --- | --- | --- | --- | --- | --- | --- | --- | --- | --- | --- |
|  | *BRCA*+ |  | *BRCA*- |  | p-Value | *BRCA*+ |  | *BRCA*- |  | p-Value |
|  | n | % | n | % |  | n | % | n | % |  |
| **Age Initial, Median (IQR)** | 60<br>(52-67) |  | 63<br>(55-69) |  | 0.154 | 62<br>(51-71) |  | 68<br>(61-74) |  | 0.002 |
| **GT Performed Pre-Initial** | 38 | 39% | 10 | 26% | 0.230 | 14 | 24% | 25 | 11% | 0.018 |
| **Initial BI-RADS** | **n=97** |  | **n=38** |  | 0.337 | **n=59** |  | **n=228** |  | <0.001 |
| 1 - Negative | 58 | 60% | 11 | 29% |  | 15 | 25% | 9 | 4% |  |
| 2 - Benign | 35 | 36% | 18 | 47% |  | 22 | 37% | 153 | 67% |  |
| 3 – Probably benign | 3 | 3% | 3 | 8% |  | 1 | 2% | 6 | 3% |  |
| 4 – Suspicious | 1 | 1% | 4 | 11% |  | 12 | 20% | 32 | 14% |  |
| 5 – Highly susp malignancy | 0 | 0% | 2 | 5% |  | 9 | 15% | 26 | 11% |  |
| No data | 0 | 0% | 0 | 0% |  | 0 | 0% | 2 | 1% |  |
| **Initial MBC Dx** | **0** | **0%** | **6** | **16%** | **N/A** | **17** | **29%** | **47** | **21%** | **0.219** |
| **Non-Baseline BI-RADS** | **n=117 (56)** |  | **n=17 (15)** |  | 0.001 | **n=16 (13)** |  | **n=40 (38)** |  | 0.049 |
| 1 - Negative | 59 | 50% | 5 | 29% |  | 4 | 25% | 2 | 5% |  |
| 2 - Benign | 57 | 49% | 10 | 59% |  | 7 | 44% | 32 | 80% |  |
| 3 – Probably benign | 0 | 0% | 2 | 12% |  | 2 | 13% | 3 | 8% |  |
| 4 – Suspicious | 1 | 1% | 0 | 0% |  | 3 | 19% | 2 | 5% |  |
| 5 – Highly susp malignancy | 0 | 0% | 0 | 0% |  | 0 | 0% | 1 | 3% |  |
| No data | 0 | 0% | 0 | 0% |  | 0 | 0% | 0 | 0% |  |
| **Non-Baseline MBC Dx** | **1** | **1%** | **0** | **0%** | **N/A** | **3** | **19%** | **4** | **10%** | **0.395** |

Abbreviations: BI-RADS, Breast Imaging Reporting and Data System; Dx, Diagnosis; GT, Genetic testing; IQR, Interquartile range; MBC, Male breast cancer

A total of 134 screening mammograms were identified that occurred after the first mammogram on record, defined as a non-baseline mammogram, and included 56 *BRCA*+ males and 15 *BRCA*-males (**Table 2, Supplemental Table 4**). Nearly all non-baseline screening-indicated examinations were categorized as BI-RADS 1-3 (*BRCA+*: 116/117, 99%; *BRCA*-: 17/17, 100%) (p=0.001) and only one (1%) MBC was identified in a *BRCA*+ male. Among symptom-indicated non-baseline mammograms, BI-RADS 4/5 findings were identified in 3/16 (19%) *BRCA*+ and 3/40 (8%) *BRCA*-mammograms (p=0.048) (**Table 2, Supplemental Table 4**). MBC was diagnosed in 3/16 (19%) *BRCA*+ males and 4/40 (10%) *BRCA*-males undergoing non-baseline symptom-indicated imaging (p=0.395).

### Diagnostic performance of mammography in BRCA+ and BRCA-males

To evaluate breast density and diagnostic performance of mammography, screening-and symptom-indicated examinations were combined. Breast density distributions were significantly different between *BRCA*+ and *BRCA*-males (p<0.001) (**Supplemental Table 5-6**), due to a higher rate of Category 3 heterogeneously dense breast tissue in *BRCA*-(10%) compared to *BRCA*+ males (1%). Among BI-RADS 4 lesions, the PPV for MBC was similar between *BRCA*+ and *BRCA*-males, with malignancy confirmed in 12/17 (71%) lesions and 28/38 (74%) lesions, respectively (p=1.000) (**Table 3**). BI-RADS 5 assessments demonstrated a high PPV in both cohorts, with cancer diagnosed in all 9 *BRCA*+ cases (100%) and 27/29 (93%) *BRCA*-cases (p=1.000) (**Table 3**).

**Table 3:** Positive Predictive Value of BI-RADS4/5 lesions detected on mammograms in males.

|  |  | **BRCA+** |  | **BRCA-** |  | **p-Value** |
| --- | --- | --- | --- | --- | --- | --- |
|  |  | n | % | n | % |  |
| **BI-RADS 4** |  | **n = 17** |  | **n = 38** |  |  |
|  | PPV | 12 | 71% | 28 | 74% | 1.000 |
| **BI-RADS 5** |  | **n = 9** |  | **n = 29** |  |  |
|  | PPV | 9 | 100% | 27 | 93% | 1.000 |

Abbreviations: BI-RADS, Breast Imaging Reporting and Data System; PPV, Positive Predictive Value

### Male breast cancer (MBC) characteristics in BRCA+ versus BRCA-males

22 *BRCA*+ and 57 *BRCA*-males were diagnosed with MBC on mammogram. The median age at diagnosis was similar between *BRCA*+ and *BRCA*-males [68 (60-75) versus 72 (63-75), p=0.25] (**Table 4, Supplemental Table 7**). Among *BRCA*+ males, 20/22 (91%) carried a *BRCA2* PGV and 2/22 (9%) carried a *BRCA1* PGV. Tumor characteristics were comparable between *BRCA*+ and *BRCA*-males. Invasive ductal carcinoma (IDC) was the predominant histology (20/22, 91% vs 53/57, 93%; p=0.669), the remainder were diagnosed with ductal carcinoma in situ (DCIS) alone. Most tumors were ER+ and HER2-(*BRCA*+, 18/22, 82% vs *BRCA*-49/57, 86%, p=0.651); HER2+ disease was identified in 14% (3/22) of *BRCA*+ and 8% (5/57) of *BRCA*-patients. Triple-negative breast cancer was uncommon in both groups (*BRCA*+, 1/22, 5% vs *BRCA*-, 1/57, 2%, respectively, p=0.651).

**Table 4:** Characteristics of Breast Cancers in *BRCA*+ vs *BRCA*-males.

|  | *BRCA*+<br>n=22 |  | *BRCA*-<br>n=57 |  | p-Value |
| --- | --- | --- | --- | --- | --- |
|  | n | % | n | % |  |
| **Age at Diagnosis** |  |  |  |  |  |
| Median (IQR) | 68 (60 – 75) |  | 72 (63 – 75) |  | 0.247 |
| **Germline mutation status** |  |  |  |  |  |
| *BRCA1* | 2 | 9% | N/A |  | N/A |
| *BRCA2* | 20 | 91% |  |  |  |
| **Histology** |  |  |  |  |  |
| Invasive carcinoma | 20 | 91% | 53 | 93% | 0.669 |
| DCIS Only | 2 | 9% | 4 | 7% |  |
| **Hormone Receptor Status** |  |  |  |  |  |
| ER+Her2- | 18 | 82% | 49 | 86% | 0.651 |
| Her2+ (ER+ or ER-) | 3 | 14% | 5 | 8% |  |
| TNBC | 1 | 5% | 1 | 2% |  |
| Other/Incomplete Data | 0 | 0% | 2 | 4% |  |
| **Node Positive/Advanced** |  |  |  |  |  |
| N0 | 10 | 45% | 32 | 56% | 0.451 |
| N1, N2, or N3 | 12 | 55% | 24 | 42% |  |
| Nx/no data | 0 | 0% | 1 | 2% |  |
| **Contralateral BC at Dx** |  |  |  |  |  |
|  | 1 | 5% | 1 | 2% | 0.482 |
| **Metastatic** |  |  |  |  |  |
| De-novo Metastatic | 3 | 14% | 4 | 7% | 0.391 |
| **Outcomes** |  |  |  |  |  |
| Local Recurrence | 1 | 5% | 1 | 2% | 0.482 |
| Subsequent Contralateral BC | 1 | 5% | 1 | 2% | 0.482 |
| Metastatic at any point | 4 | 18% | 9 | 16% | 0.748 |
| **Oncotype DX genomic test Recurrence Score\*** | n = 9 |  | n=31 |  |  |
| Low (0-11) | 1 | 11% | 7 | 23% | 0.510 |
| Intermediate (12-25) | 3 | 33% | 7 | 23% |  |
| High (>25) | 2 | 22% | 3 | 10% |  |
| Not Completed | 3 | 33% | 14 | 45% |  |

\*Oncotype DX genomic test Recurrence Score reported only for ER+, N0 patients
Abbreviations: BC, Breast Cancer; DCIS, Ductal carcinoma in-situ; Dx, Diagnosis; ER, Estrogen Receptor; HER2, Human Epidermal growth factor Receptor 2; IQR, Interquartile Range; TNBC, Triple Negative Breast Cancer

At diagnosis, node-positive disease (N1-N3) was observed in 55% (12/22) of *BRCA*+ and 42% (24/57) of *BRCA*-males (p=0.451). Among ER+, node-negative patients with available Oncotype DX genomic testing, high risk scores (>25) were observed in 22% (2/9) of *BRCA*+ compared with 10% (3/31) of *BRCA*-males, whereas low-risk scores (0-11) were identified in 11% (1/9) of *BRCA+* males compared with 23% (7/31) of *BRCA-*males (p=0.510).

Rates of synchronous contralateral breast cancer were low and did not differ significantly between *BRCA*+ and *BRCA*-males (1/22, 5% vs 1/57, 2%, p=0.482). The frequency of de novo metastatic disease was also similar between *BRCA*+ and *BRCA*-males (3/22, 14% vs 4/57, 7%, p=0.391). Both *BRCA*+ and *BRCA*-males had low rates of subsequent contralateral breast cancer (1/22, 5% vs 1/57, 2%, p=0.482). While local recurrence was rare (1/22, 5% vs 1/57, 2%, p=0.482), both *BRCA*+ and *BRCA*-males had high rates of having metastatic disease at any time in their disease course (4/22, 18% vs 9/57, 16%, p=0.748).

## Discussion

Male breast cancer is a rare disease, but the risk is substantially increased among males with pathogenic *BRCA1/2* variants^9,12–16^. NCCN guidelines currently state that consideration be given to annual screening mammography starting at age 50, particularly for males with *BRCA2* PV^20,21^. The current guideline is based on limited evidence derived primarily from retrospective studies and small institutional cohorts, reflecting the broader lack of prospective data in male breast cancer screening^2,9,22,24,28^. Consideration of annual mammography was added to NCCN guidelines in February 2023. In this multi-institutional cohort of 810 *BRCA*+ males; screening-indicated mammography was infrequently utilized despite most patients being age-eligible for screening. We identified gaps in screening-related documentation and care coordination and found that male breast cancers were more commonly detected following symptom-indicated compared to screening-indicated mammography.

An important observation from this study was the low rate of screening mammography among *BRCA*+ males. The NCCN added the comment that annual screening mammography should be considered for *BRCA+* males in 2023. Even though 60% of discussions occurred after this addition, 84% of age-eligible males had no documentation of prior mammography. These findings are consistent with prior reports demonstrating limited adoption of breast cancer screening among high-risk men^23,24^. Men with *BRCA* mutations frequently report uncertainty regarding their breast cancer risk and available screening recommendations^23^, and have reported discomfort related to perceived female-centered breast imaging environment^29^. Our findings suggest that these challenges persist even within large academic and integrated healthcare systems.

Among patients who underwent mammography, screening-indicated examinations demonstrated a low diagnostic yield. Nearly all screening studies were categorized as BI-RADS 1-3 and yielded few suspicious findings or cancer diagnoses in both *BRCA*+ and *BRCA*-males. In contrast, symptom-indicated imaging was associated with substantially higher rates of suspicious findings and cancer diagnoses. Nearly one-third (29%) of *BRCA*+ males undergoing a first symptom-indicated imaging were diagnosed with MBC. This observation mirrors prior high-risk screening cohorts, which have reported low uptake of asymptomatic screening-indicated imaging, with high-risk men more frequently presenting with symptoms^22,24^. Our findings should not be interpreted as evidence that screening mammography lacks value in *BRCA*+ males. Rather, they suggest that existing screening recommendations may not be reaching enough eligible patients to meaningfully affect patterns of cancer detection. Interestingly, MBC diagnosis rates were similar between *BRCA+* and *BRCA*-males. This finding underscores a potential missed opportunity for identification of other high-risk men.

The positive predictive value (PPV) of BI-RADS 4 and 5 findings was high in both *BRCA*+ and *BRCA*-males. Malignancy was confirmed in 71% of BI-RADS 4 lesions and nearly all BI-RADS 5 lesions. In a prior retrospective study of 75 men with suspicious breast findings, mammographic PPV was 87% for BI-RADS 4/5 lesions^28^. Notably, the PPVs observed in our cohort exceeded those typically reported in women, where BI-RADS 4 lesions are associated with PPVs of approximately 8%, 22%, and 69% for BI-RADS 4A, 4B and 4C lesions, respectively^25–27^. The higher PPVs observed in men likely reflect the lower prevalence of benign breast lesions and use of mammography mainly in the setting of symptoms rather than routine population-based screening. *BRCA* status did not influence the predictive value of BI-RADS 4/5 findings, suggesting that suspicious mammographic lesions warrant a similar level of clinical concern in males regardless of underlying genetic risk.

The clinicopathologic characteristics of MBC were largely similar between *BRCA+* and *BRCA-*male patients in our cohort. Consistent with prior studies, most cancers occurred in *BRCA2+* carriers^9,12–16^. Tumors in both groups were overwhelmingly hormone-receptor positive invasive carcinomas, mirroring the established biology of MBC^4^. *BRCA+* males demonstrated marginally higher rates of node-positive and de novo metastatic disease compared to *BRCA*-males. Among ER+ node-negative patients who underwent Oncotype DX genomic testing, there was a suggestion that *BRCA*+ males were more likely to have high risk recurrence scores, consistent with prior reports suggesting that *BRCA*-associated tumors may exhibit more aggressive features^2,9^.

Several limitations should be considered when interpreting these findings. As a retrospective study, our analysis was dependent on the completeness and accuracy of electronic health record documentation, which may have resulted in under ascertainment of screening discussions, mammography utilization, and genetic testing information. Imaging performed outside the Penn or VA health systems may not have been captured, and classification of imaging indication relied on clinical documentation that may be incomplete or misclassified. The relatively small number of MBC diagnoses, particularly among screening-indicated examinations, limited our ability to evaluate screening effectiveness. Additionally, because current NCCN guidelines state that *BRCA+* males can consider mammography rather than recommending it, this ambiguous language can complicate clinical implementation efforts to define and measure adequate screening. Despite these limitations, this study represents the largest evaluation of mammography utilization among *BRCA+* males and provides valuable insight into real-world screening practices among *BRCA+* males.

In conclusion, mammography remains underutilized among *BRCA*+ males despite elevated hereditary risk and existing guidelines for the consideration of mammograms. Gaps in care coordination and implementation of risk-based screening may contribute to missed opportunities for earlier cancer detection as both *BRCA*+ and *BRCA*-males are diagnosed with MBC at more advanced stages. These findings highlight the need for improved integration of high-risk cancer screening into routine clinical practice and underscore the importance of prospective studies to better define the role of mammography (including when to start, and the frequency of the exams) in high-risk male populations.

## Data Availability

All data produced in the present work are contained in the manuscript.

## Acknowledgements

This work was supported in part by the United States (U.S.) Department of Veterans Affairs Office of Research and Development (1I50CU000193, KWR; 1I01CX002710, MWS, KNM, IPG), National Cancer Institute (P30CA016520, PEG, KLN), Prostate Cancer Foundation (18VALO08, KWR; 22CHAL02, KNM, IPG), the National Center for Advancing Translational Sciences (KL2TR001856, EMB), American Cancer Society (RSG-23-1038098-01-HOPS, AMM), the Center of Innovation to Accelerate Discovery and Practice Transformation at the Durham VA Health Care System (CIN 13-410, LLZ), the Basser Center for *BRCA* (SMD, JQZ, KNM), VA National Oncology Program Pre-Career Development Award Program (EMB), and VA National Oncology Program (MJK). The contents do not represent the views of the U.S. Department of Veterans Affairs or the United States Government.

