## Supplemental Tables for "Mammography utilization and male breast cancer in *BRCA1/2* carriers"

**Supplementary Tables**

**Supplemental Table 1: Documentation of Mammogram Discussions in *BRCA*+ Mammography Decision Making Cohort (Males Age ≥ 50 Years)**

| | **Penn n = 322** | | **VA n = 240** | | **Combined n = 562** | |
|---|---|---|---|---|---|---|
| | n | % | n | % | n | % |
| *No discussion documented* | 142 | 44% | 65 | 27% | 207 | 37% |
| *Discussed by a genetic counselor but no ordering provider discussion* | 25 | 8% | 82 | 34% | 107 | 19% |
| *SDM discussion against mammogram* | 82 | 25% | 52 | 22% | 134 | 24% |
| *SDM for a mammogram but not completed* | 17 | 5% | 6 | 3% | 23 | 4% |
| *Completed mammogram* | 56 | 17% | 35 | 15% | 91 | 16% |

Abbreviations: SDM, shared decision-making; VA, Veterans Health Administration

**Supplemental Table 2: Demographics of Mammogram/MBC Cohort stratified by site**

| | **Penn *BRCA+* n=85** | | **VA *BRCA+* n=67** | | **Penn *BRCA-* n=66** | | **VA *BRCA-* n=200** | |
|---|---|---|---|---|---|---|---|---|
| | n | % | n | % | n | % | n | % |
| ***Self-Identified Race*** | | | | | | | | |
| *White* | 77 | 91% | 50 | 75% | 54 | 82% | 129 | 65% |
| *Black or African American* | 3 | 4% | 11 | 16% | 8 | 12% | 59 | 30% |
| *Asian* | 0 | 0% | 1 | 1% | 1 | 2% | 2 | 1% |
| *Multiple Races* | 0 | 0% | 0 | 0% | 3 | 5% | 0 | 0% |
| *Other/Unknown* | 5 | 6% | 5 | 7% | 0 | 0% | 10 | 5% |
| ***Ethnicity*** | | | | | | | | |
| *Non-Hispanic/Latino* | 84 | 99% | 57 | 85% | 66 | 100% | 174 | 87% |
| *Hispanic* | 1 | 1% | 6 | 9% | 0 | 0% | 15 | 8% |
| *Unknown* | 0 | 0% | 4 | 6% | 0 | 0% | 11 | 6% |
| ***Genetic Testing*** | | | | | | | | |
| *BRCA1+* | 23 | 27% | 21 | 31% | NA | NA | NA | NA |
| *BRCA2+* | 61 | 72% | 46 | 69% | NA | NA | NA | NA |
| *BRCA1+ & BRCA2+* | 1 | 1% | 0 | 0% | NA | NA | NA | NA |

Abbreviations: MBC, Male breast cancer; NA, Not Applicable; VA, Veterans Health Administration

**Supplemental Table 3: Results of Overall BI-RADS Categories stratified by site**

| | Screening Indicated Mammograms | | | | | | | | | |
|---|---|---|---|---|---|---|---|---|---|---|
| | Penn *BRCA+* | | Penn *BRCA-* | | p - Value | VA *BRCA+* | | VA *BRCA-* | | p - Value |
| | n | % | n | % | | n | % | n | % | |
| **Age Initial Mammogram Completed, Median (IQR)** | 58 (51 - 64) | | 67 (62 - 73) | | 0.113 | 62 (54 - 70) | | 62 (55 - 69) | | 0.966 |
| **Initial BI-RADS** | **n=58** | | **n=10** | | 0.001 | **n=39** | | **n=28** | | 0.184 |
| 1 - Negative | 39 | 67% | 4 | 40% | | 19 | 49% | 7 | 25% | |
| 2 - Benign | 17 | 29% | 2 | 20% | | 18 | 46% | 16 | 57% | |
| 3 - Probably benign | 1 | 2% | 0 | 0% | | 2 | 5% | 3 | 11% | |
| 4 - Suspicious | 1 | 2% | 3 | 30% | | 0 | 0% | 1 | 4% | |
| 5 – High susp malignancy | 0 | 0% | 1 | 10% | | 0 | 0% | 1 | 4% | |
| No data | 0 | 0% | 0 | 0% | | 0 | 0% | 0 | 0% | |
| **Initial MBC Dx** | 0 | 0% | 4 | 40% | N/A | 0 | 0% | 2 | 7% | 0.171 |
| **Non-Baseline BI-RADS** | **n=96 (41)** | | **n=2 (2)** | | 0.930 | **n=21 (15)** | | **n=15 (13)** | | 0.276 |
| 1 - Negative | 51 | 53% | 1 | 50% | | 8 | 38% | 4 | 27% | |
| 2 - Benign | 45 | 47% | 1 | 50% | | 12 | 57% | 9 | 60% | |
| 3 - Probably benign | 0 | 0% | 0 | 0% | | 0 | 0% | 2 | 13% | |
| 4 - Suspicious | 0 | 0% | 0 | 0% | | 1 | 5% | 0 | 0% | |
| 5 – High susp malignancy | 0 | 0% | 0 | 0% | | 0 | 0% | 0 | 0% | |
| No data | 0 | 0% | 0 | 0% | | 0 | 0% | 0 | 0% | |
| **Non-Baseline MBC Dx** | 0 | 0% | 0 | 0% | N/A | 1 | 5% | 0 | 0% | N/A |

| | Symptom Indicated Mammograms | | | | | | | | | |
|---|---|---|---|---|---|---|---|---|---|---|
| | Penn *BRCA+* | | Penn *BRCA-* | | p - Value | VA *BRCA+* | | VA *BRCA-* | | p - Value |
| | n | % | n | % | | n | % | n | % | |
| **Age Initial Mammogram Completed, Median (IQR)** | 59 (47 - 65) | | 67 (57 - 74) | | 0.220 | 63 (56 - 74) | | 75 (66 - 79) | | 0.091 |
| **Initial BI-RADS** | **n=31** | | **n=56** | | 0.078 | **n=28** | | **n=172** | | <0.001 |
| 1 - Negative | 9 | 29% | 5 | 9% | | 6 | 21% | 4 | 2% | |
| 2 - Benign | 11 | 35% | 35 | 63% | | 11 | 39% | 118 | 69% | |
| 3 - Probably benign | 1 | 3% | 2 | 4% | | 0 | 0% | 4 | 2% | |
| 4 - Suspicious | 6 | 19% | 7 | 13% | | 6 | 21% | 25 | 15% | |
| 5 – High susp malignancy | 4 | 13% | 7 | 13% | | 5 | 18% | 19 | 11% | |
| No data | 0 | 0% | 0 | 0% | | 0 | 0% | 2 | 1% | |
| **Initial MBC Dx** | 9 | 30% | 9 | 16% | 0.397 | 9 | 32% | 38 | 22% | 0.240 |
| **Non-Baseline BI-RADS** | **n=14 (11)** | | **n=6 (6)** | | 0.827 | **n=2 (2)** | | **n=34 (32)** | | 0.002 |
| 1 - Negative | 4 | 29% | 1 | 17% | | 0 | 0% | 1 | 3% | |
| 2 - Benign | 7 | 50% | 4 | 67% | | 0 | 0% | 28 | 82% | |
| 3 - Probably benign | 2 | 14% | 1 | 17% | | 0 | 0% | 2 | 6% | |
| 4 - Suspicious | 1 | 7% | 0 | 0% | | 2 | 100% | 2 | 6% | |
| 5 – High susp malignancy | 0 | 0% | 0 | 0% | | 0 | 0% | 1 | 3% | |
| No data | 0 | 0% | 0 | 0% | | 0 | 0% | 0 | 0% | |
| **Non-Baseline MBC Dx** | 1 | 7% | 1 | 17% | 0.521 | 2 | 100% | 3 | 9% | 0.018 |

Abbreviations: BI-RADS, Breast Imaging Reporting and Data System; IQR, Interquartile range; VA, Veterans Health Administration

**Supplemental Table 4: Results of BI-RADS Density Categories in *BRCA*+ versus *BRCA*- Males**

| | ***BRCA*+ n=289 (155)** | | ***BRCA*- n=323 (266)** | | **p-value** |
|---|---|---|---|---|---|
| | n | % | n | % | <0.001 |
| 1 – Breasts are almost entirely fatty | 165 | 57% | 152 | 47% | |
| 2 – scattered fibroglandular density | 79 | 27% | 112 | 35% | |
| 3 – Heterogeneously dense | 4 | 1% | 32 | 10% | |
| 4 – Extremely dense | 0 | 0% | 1 | 0% | |
| No data | 41 | 14% | 26 | 8% | |

Abbreviations: BI-RADS, Breast Imaging Reporting and Data System

**Supplemental Table 5: Results of BI-RADS Density Categories stratified by site and mammogram instance**

| | Screening Indicated Mammograms | | | | | | | | | | Symptom Indicated Mammograms | | | | | | | | | |
|---|---|---|---|---|---|---|---|---|---|---|---|---|---|---|---|---|---|---|---|---|
| | **Penn *BRCA+*** | | **Penn *BRCA-*** | | **p - Value** | **VA *BRCA+*** | | **VA *BRCA-*** | | **p - Value** | **Penn *BRCA+*** | | **Penn *BRCA-*** | | **p - Value** | **VA *BRCA+*** | | **VA *BRCA-*** | | **p - Value** |
| | **n** | **%** | **n** | **%** | | **n** | **%** | **n** | **%** | | **n** | **%** | **n** | **%** | | **n** | **%** | **n** | **%** | |
| **Initial BI-RADS Density** | **n = 58** | | **n = 10** | | 0.030 | **n = 39** | | **n = 28** | | 0.029 | **n = 31** | | **n = 56** | | 0.239 | **n = 28** | | **n = 172** | | 0.913 |
| 1 - Breasts are almost entirely fatty | 41 | 71% | 6 | 60% | | 25 | 64% | 14 | 50% | | 12 | 39% | 37 | 66% | | 13 | 46% | 75 | 44% | |
| 2 - Scattered areas of fibroglandular density | 6 | 10% | 3 | 30% | | 12 | 31% | 5 | 18% | | 10 | 32% | 16 | 29% | | 10 | 36% | 64 | 37% | |
| 3 - Heterogeneously dense | 0 | 0% | 1 | 10% | | 0 | 0% | 4 | 14% | | 0 | 0% | 3 | 5% | | 2 | 7% | 16 | 9% | |
| 4 - Extremely dense | 0 | 0% | 0 | 0% | | 0 | 0% | 0 | 0% | | 0 | 0% | 0 | 0% | | 0 | 0% | 0 | 0% | |
| No data | 11 | 19% | 0 | 0% | | 2 | 5% | 5 | 18% | | 9 | 29% | 0 | 0% | | 3 | 11% | 17 | 10% | |
| **Non-baseline BI-RADS Density** | **n=96 (41)** | | **n=2 (2)** | | 0.530 | **n=21 (15)** | | **n=15 (13)** | | 0.082 | **n=14 (11)** | | **n=6 (6)** | | 0.125 | **n=2 (2)** | | **n=34 (32)** | | 0.023 |
| 1 - Breasts are almost entirely fatty | 58 | 60% | 1 | 50% | | 8 | 38% | 6 | 40% | | 8 | 57% | 4 | 67% | | 0 | 0% | 9 | 26% | |
| 2 - Scattered areas of fibroglandular density | 24 | 25% | 1 | 50% | | 13 | 62% | 6 | 40% | | 4 | 29% | 0 | 0% | | 0 | 0% | 17 | 50% | |
| 3 - Heterogeneously dense | 0 | 0% | 0 | 0% | | 0 | 0% | 3 | 20% | | 0 | 0% | 1 | 17% | | 2 | 100% | 4 | 12% | |
| 4 - Extremely dense | 0 | 0% | 0 | 0% | | 0 | 0% | 0 | 0% | | 0 | 0% | 0 | 0% | | 0 | 0% | 1 | 3% | |
| No data | 14 | 15% | 0 | 0% | | 0 | 0% | 0 | 0% | | 2 | 14% | 1 | 17% | | 0 | 0% | 3 | 9% | |

Abbreviations: BI-RADS, Breast Imaging Reporting and Data System; VA, Veterans Health Administration

**Supplemental Table 6: Characteristics of Male Breast Cancer stratified by site**

| | Penn *BRCA*+ n=10 | | Penn *BRCA*- n=14 | | VA *BRCA*+ n=12 | | VA *BRCA*- n=43 | |
|---|---|---|---|---|---|---|---|---|
| | n | % | n | % | n | % | n | % |
| **Age at Diagnosis** | | | | | | | | |
| Median (IQR) | 61 (55 – 68) | | 64 (56-72) | | 73 (65-75) | | 73 (64-77) | |
| **Germline mutation status** | | | | | | | | |
| BRCA1 | 0 | 0% | N/A | | 2 | 17% | N/A | |
| BRCA2 | 10 | 100% | | | 10 | 83% | | |
| **Histology** | | | | | | | | |
| Invasive carcinoma | 9 | 90% | 13 | 93% | 11 | 92% | 40 | 93% |
| DCIS Only | 1 | 10% | 1 | 7% | 1 | 8% | 3 | 7% |
| Unknown/Other | 0 | 0% | 0 | 0% | 0 | 0% | 0 | 0% |
| **Hormone Receptor Status** | | | | | | | | |
| ER+HER2- | 9 | 90% | 13 | 93% | 9 | 75% | 36 | 84% |
| HER2+ (ER+ or ER-) | 1 | 10% | 1 | 7% | 2 | 17% | 4 | 9% |
| TNBC | 0 | 0% | 0 | 0% | 1 | 8% | 1 | 2% |
| Other/Incomplete Data | 0 | 0% | 0 | 0% | 0 | 0% | 2 | 5% |
| **Node Positive/Advanced** | | | | | | | | |
| N0 | 3 | 30% | 6 | 43% | 7 | 58% | 26 | 60% |
| N1, N2, or N3 | 7 | 70% | 7 | 50% | 5 | 42% | 17 | 40% |
| Nx/no data | 0 | 0% | 1 | 7% | 0 | 0% | 0 | 0% |
| **Contralateral BC at Dx** | | | | | | | | |
| | 0 | 0% | 1 | 7% | 1 | 8% | 0 | 0% |
| **Metastatic** | | | | | | | | |
| De novo Metastatic | 2 | 20% | 0 | 0% | 1 | 8% | 4 | 9% |
| **Outcomes** | | | | | | | | |
| Local Recurrence | 1 | 10% | 0 | 0% | 0 | 0% | 1 | 2% |
| Subsequent Contralateral BC | 0 | 0% | 0 | 0% | 1 | 8% | 1 | 2% |
| Metastatic at any time | 3 | 30% | 1 | 7% | 1 | 8% | 7 | 16% |
| **Oncotype Dx genomic test Recurrence Score*** | n=3 | | n=6 | | n=6 | | n=25 | |
| Low (0-11) | 1 | 33% | 0 | 0% | 0 | 0% | 7 | 28% |
| Intermediate (12-25) | 2 | 66% | 0 | 0% | 1 | 17% | 7 | 28% |
| High (>25) | 0 | 0% | 0 | 0% | 3 | 33% | 3 | 12% |
| Not Complete | 0 | 0% | 6 | 100% | 3 | 50% | 8 | 32% |

*Oncotype Recurrence Score reported only for ER+, N0 patients
